# Discrepancies in the detection of SARS-CoV-2 by qRT-PCR are dependent on the target gene used for its amplification: Implications in the diagnosis of clinical infection

**DOI:** 10.1101/2021.08.30.21262536

**Authors:** Elena Campos-Pardos, Juan Calvet-Seral, Antonio Aguilera, Ana Milagro-Beamonte, Ana Martínez-Sapiña, Gema Barbeito, María Luisa Pérez del Molino, Jesús Gonzalo-Asensio

## Abstract

Discrepancies exist in Cycle threshold (Ct) values during detection of SARS-CoV-2 by qRT-PCR. We demonstrate that Ct values depend on the position of the target gene in the viral genome. Simultaneous detection of five genes in positive samples revealed lower Ct values as we move further to the 3’ end (orf1AB/RdRp>E>M>orf7a>N). These findings were confirmed in a retrospective analysis with 363 positive clinical samples. Our findings have key implications in clinical diagnostics of SARS-CoV-2, patient management and public health interventions.

## Introduction

Detection of Severe Acute Respiratory Syndrome CoronaVirus 2 (SARS-CoV-2) by quantitative Reverse Transcription Polymerase Chain Reaction (qRT-PCR) tests usually relies in considering sample positivity when the Cycle Threshold (Ct) Value is below a standardized value (i.e. Ct cutoff=34 or 35). However, this method does not necessarily provide information about viral replication, viral load or disease transmissibility [1, 2]. In addition, after more than one year of the beginning of the COronaVIrus Disease pandemic of 2019 (COVID-19), there is a plethora of commercial kits which detects different genomic regions of the SARS-CoV-2 RNA, each with different sensitivities [3]. However, an aspect completely overlooked in qRT-PCR detection of SARS-CoV-2 is inherently related to the own biology of betacoronaviruses. This virus genus is characterized by large viral genomes, which are discontinuously transcribed in form of subgenomic mRNAs [4]. Specifically, the SARS-CoV-2 original strain [5] contains up to 10 subgenomic mRNAs characterized by containing the same 3’ ends as the genomic RNA, but different 5’ ends which contains the start codon of each downstream gene (Figure 1A). Accordingly, RNAs of genes closer to the 3’ end will contain their own subgenomic mRNAs, as well as subgenomic mRNAs from upstream genes and the RNA molecule from the own viral genome (Figure 1A). This transcriptome architecture might have key implications in the qRT-PCR diagnostic of SARS-CoV-2 due to the increased accumulation of viral transcripts as we progress in the viral genome as was previously demonstrated by RNA sequencing [6, 7]. Specifically, we hypothesized that Ct values within a given sample would gradually decrease as we move further to the 3’ end of the SARS-CoV-2 genome, due to the intrinsic transcriptome of the SARS-CoV-2

**Figure 1 legend.**
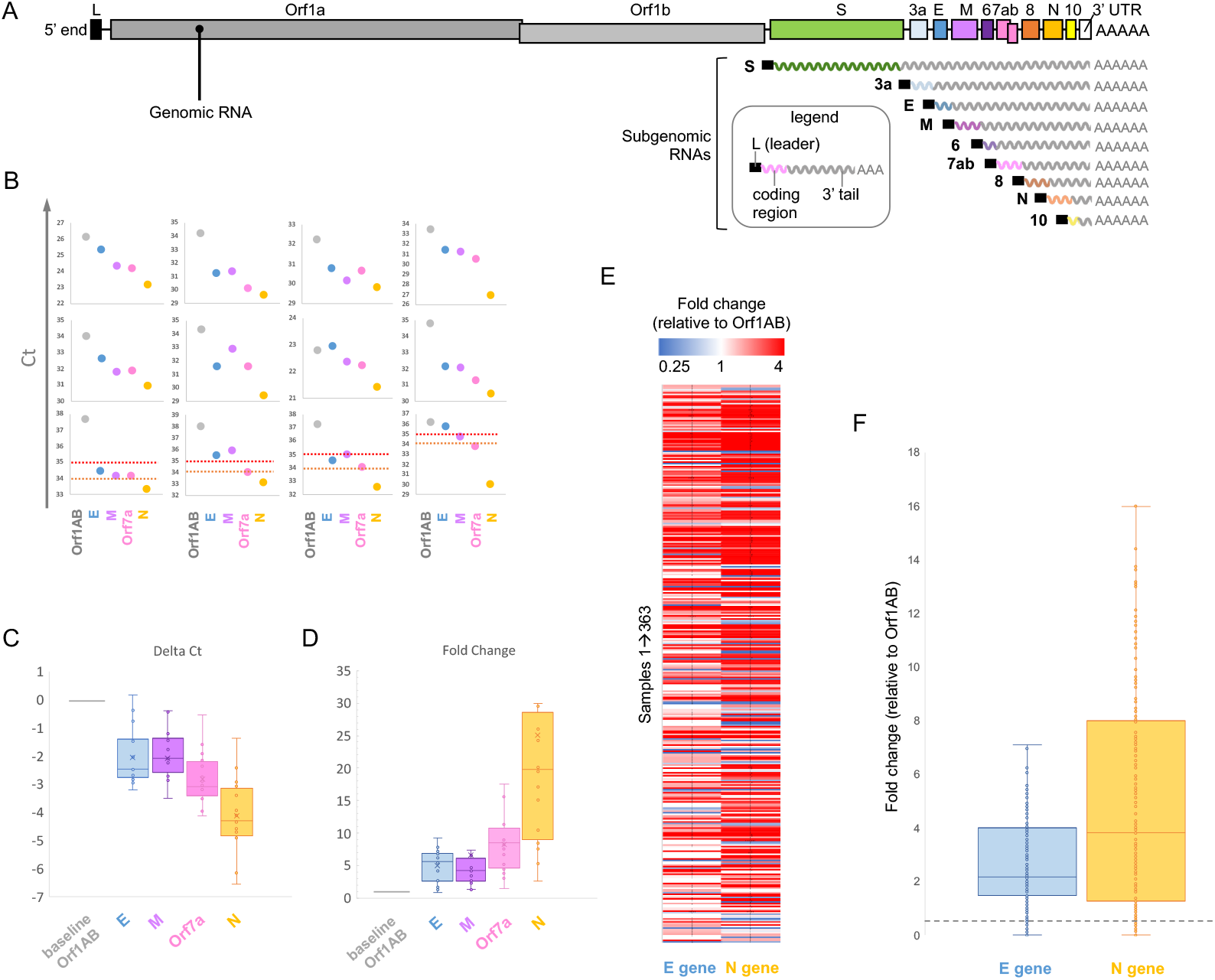
**A**. SARS-CoV-2 genome and transcriptome organization. The transcripts expressed as subgenomic mRNAs are indicated. Note the enrichment in viral RNA as genes are positioned closer to the 3’ end. Also note the position of the main target genes used in qRT-PCR diagnostics (orf1ab, E, M and N) relative to the 3’ end of the viral genome. **B**. Ct values of orf1ab, E, M, orf7a and N genes in clinical samples previously validates as positive. Ct cutoffs of 34 and 35 are indicated in the 4 lower individual panels by orange and red lines, respectively. **C**. Box plots showing Delta Ct values of E, M, orf7a and N genes relative to the orf1ab. **D**. Box plots showing fold change in gene expression of E, M, orf7a and N genes relative to the orf1ab. **E**. Heat map showing fold change in E and N gene expression relative to orf1ab in 363 positive clinical samples. **F**. Box plot showing fold change in E and N gene expression relative to orf1ab in samples used for the retrospective analysis. The gray discontinuous line indicates the orf1ab baseline. Note the gradual increase in gene expression for E and N genes.

### Study design

We performed a two-step characterization of Ct values in SARS-CoV-2 positive clinical samples obtained from oro-pharyngeal swabs. On the one hand, we simultaneously analysed the expression of orf1AB (RdRp), E, M, orf7a and N genes in a subset of samples to characterize the viral transcriptome in terms of Ct values of genes positioned across the viral genome. On the other hand, we performed a retrospective analysis of 363 positive samples in which orf1AB, E and N genes were detected.

## Results

### Transcriptomic characterization of SARS-CoV-2 positive samples reveal progressively lower Ct values of genes more proximal to the 3’ end of the viral genome

Sixteen SARS-CoV-2 RNA positive samples were reverse transcribed and used for qRT-PCR detection of orf1AB, E, M, orf7a and N genes. Each gene within a specific sample were detected in triplicate and the average Ct was calculated. Results demonstrated an overall progressive reduction of Ct values in genes positioned closest to the 3’ end of the viral genome. Accordingly, we found the following trend in Ct values: orf1AB>E>M>orf7a>N genes (Figure 1B). As expected, the different samples differed in their Ct values, which is a putative indicator of diverse viral RNA loads between samples, and demonstrate that our results can be extrapolated to different clinical scenarios. Our findings are especially relevant for those clinical samples showing high Ct values. As exemplified in the 4 lower panels of Figure 1B, Ct cutoffs of 34 and 35 would result in false negative diagnostics depending on the target gene used in qRT-PCR. In order to normalize Ct values between samples, we also calculated Delta Ct values relative to Ct of the orf1AB (Ct target gene – Ct orf1AB), being this later the first gene of the SARS-CoV-2 genome which is not transcribed as subgenomic mRNAs. A box-plot analyses indicated the gradual decrease in Ct values for genes more distant from the 5’ end (Figure 1C), which indicates that our results are reproducible irrespective of the viral RNA amount present in the sample. Finally, we analyzed the fold change in gene expression for every interrogated gene relative to the orf1ab (2e-Delta Ct). Results showed a progressive enrichment in RNA as we move forward to the 3’ end (Figure 1D) and corroborate previous findings using RNA sequencing [6, 7]. Altogether, the N gene, positioned closest to the 3’ end of the viral genome, demonstrated the highest expression (Figure 1D), and consequently, the lowest Ct values (Figure 1B and C), compared to neighboring genes. Accordingly, these robust results during detection of the N gene provide advice about the suitability of this gene for high sensitive detection of SARS-CoV-2 in clinical samples.

### Retrospective analyses with SARS-CoV-2 positive samples analyzed for orf1ab, E and N genes demonstrate higher expression, and lower Ct values, in genes positioned in the 3’ end of the viral genome

To gain insight into the clinical relevance of our findings, we decided to perform a retrospective study with SARS-CoV-2 positive samples in which the orf1ab, E and N genes were detected. As expected, samples showed different Ct ranges which is putatively related to variations in viral RNA amounts between samples. Accordingly, due to this sample heterogenicity, we normalized data by calculating Delta Ct values relative to the orf1ab genes. Subsequently, we calculated the corresponding fold change in gene expression of E and N genes relative to the orf1ab genes. Individual fold changes can be visualized in the heat-map shown in Figure 1E. A global analysis of fold changes can be visualized in a box plot in Figure 1F. Notably, expression of both, E and N genes, were higher than the orf1ab. Further, E and N genes showed expression differences, being the N gene more expressed than the E gene (Figure 1E and F). Accordingly, our retrospective study remarkably correlates with transcriptomics characterization of SARS-CoV-2 by RNA sequencing [6, 7] and our own targeted study (Figure 1C), and confirms progressively higher expression of orf1ab, E and N genes, which correlate with progressively decreasing Ct values, respectively.

## Discussion

After more than one year of the COVID-19 pandemics a plethora of diagnostic methods have been developed, most of them based on qRT-PCR analyses of viral RNA, and other different techniques based on RT-LAMP or CRISPR [8]. Notably, the sensitivity of these methods not only depends on the RNA viral load in a specific clinical sample, but it also depends on the target gene used for diagnostics. As reported here and in previous transcriptomic studies of SARS-CoV-2 [6, 7], transcription of the viral genome in the form of subgenomic mRNAs results in a progressive accumulation of transcripts as we approach to the 3’ end of the genome. This inherent characteristic of the betacoronaviruses transcriptome could undoubtedly have an impact on a gene-dependent detection of Ct values used in diagnostics. Here, we confirm this hypothesis in a targeted trancriptomic study of SARS-CoV-2 positive samples which is further confirmed with a retrospective analysis of 363 clinical positive specimens. Our results have important implications for the clinical diagnostics. First: genes proximal to the 5’ end confer a higher Ct value than those genes localized in the 3’ end of the virus genome, being their detection by qRT-PCR less sensitive (Figure 1B). Second: the N gene is robustly higher expressed in positive samples, and consequently Ct values of the N gene are considerably lower relative to the remaining genes, being the most sensitive target gene for the detection of SARS-COV2 by qRT-PCR. Third: Ct values in the range of 34-35 for the N gene might correlate with either a pre-symptomatic state indicative of early viral replication or with a post-symptomatic state indicative of disease resolution. Accordingly caution should be taken considering the Ct values of the N gene and additional parameters regarding the patient clinical status or the disease prevalence should be considered.

## Conclusions

Our study highlights the importance of the target gene that is selected for the detection of SARS-CoV-2 by qRT-PCR. We provide a rationale interpretation of Ct values, which is based on the own viral biology. Results reported herein may help in both, the diagnostic interpretation, and the management of patient by public health measures. Our results provide advice about the need for standardization in molecular diagnosis of COVID-19 and postulate a biological explanation for the discrepancies observed in Ct values during qRT-PCR assays.

## Data Availability

All the published data will be available upon request

## Conflict of interest

None declared

## Funding statement

This work was supported by grants from Oficina de Transferencia de Resultados de Investigación-University of Zaragoza (reference Valle de la Muerte 2020) and by Santander-UNIZAR COVID-19 call to J. G.-A. E. C.-P. is beneficiary from a FPU grant (reference FPU17/02909) from the Spanish Ministry of Education, Culture and Sport. J. C.-S. is beneficiary from a DGA grant from Gobierno de Aragón.

## Notes

### Competing Interest Statement

The authors have declared no competing interest.

### Funding Statement

This work was supported by grants from Oficina de Transferencia de Resultados de Investigacion-University of Zaragoza (reference Valle de la Muerte 2020) and by Santander-UNIZAR COVID-19 call to J. G.-A. E. C.-P. is beneficiary from a FPU grant (reference FPU17/02909) from the Spanish Ministry of Education, Culture and Sport. J. C.-S. is beneficiary from a DGA grant from Gobierno de Aragon.

### Author Declarations

This project was evaluated by the Ethics Committee for Research in the Autonomous Community of Aragon (Comite de Etica para la Investigacion en la Comunidad Autonoma de Aragon)-CEICA (Reference SA21-12), and the ethics approval was favorable.

